# Pyelonephritis is associated with persistent alterations in immune-stromal and vascular programs in the human placenta

**DOI:** 10.64898/2026.10.05.26364820

**Authors:** Christine A. Blauvelt, Nida Ozarslan, Stephanie M. Tsoi, Olena Trush, Chloe Taylor, Sirirak Buarpung, Megan Chidboy, Jeanine S. Esteban, Amayrani Morales, Corina A. Mong, Neela Rahseparian, Cynthia Gasper, Joshua F. Robinson, Elizabeth E. Crouch, Stephanie L. Gaw

**Author notes:** **Corresponding author:** Christine A. Blauvelt - Division of Maternal-Fetal Medicine, Department of Obstetrics, Gynecology, and Reproductive Sciences, University of California San Francisco, 490 Illinois Street, 10th Floor, San Francisco, CA 94158.

## Abstract

Maternal systemic infection during pregnancy is associated with placental dysfunction even when pathogens do not directly invade the placenta, but the cellular pathways linking antecedent infection to persistent placental alterations remain poorly defined. Here we used spatial transcriptomics to resolve trophoblast-, fetal endothelial, and Hofbauer cell-enriched niches in placentas collected at term after clinical recovery from antepartum pyelonephritis. We profiled 168 spatially defined areas from 14 placentas and integrated transcriptional findings with blinded histopathology and quantitative extracellular matrix analysis. Pyelonephritis was associated with strikingly compartment-specific transcriptional responses. Hofbauer cell-enriched niches showed the most extensive alterations, with coordinated enrichment of extracellular matrix remodeling and vascular developmental programs, including increased *ELN* expression (log_2_FC = 0.84, FDR = 0.033) and increased expression of genes governing collagen deposition, elastic-fiber assembly, matrix maturation and turnover. Quantitative analysis of 1,790 fetal vessels independently identified altered perivascular matrix composition, with Hofbauer cell-enriched *ELN* expression correlating with greater perivascular elastin content (ρ = 0.67, *P* = 0.008). Fetal endothelial-enriched regions exhibited complementary alterations in vascular remodeling and endothelial identity, including increased *PLAC8* (log_2_FC = 1.36, FDR = 0.047), whereas trophoblast-enriched regions showed little detectable transcriptional response. Histopathology further demonstrated increased lesions associated with maternal vascular malperfusion and perivillous fibrin deposition. Together, these findings identify the villous immune–stromal–vascular microenvironment as a persistent site of placental remodeling following maternal infection and suggest that clinical recovery from maternal illness may not coincide with normalization of placental biology.

## Introduction

The human placenta is a highly specialized fetomaternal interface composed of a branching villous tree that facilitates nutrient exchange, gas transfer, immune regulation, and endocrine signaling between the mother and fetus. Placental development is a dynamic process that spans the entirety of gestation and is tightly coordinated with the maternal immune system.^1^

Following implantation, extravillous trophoblasts colonize and remodel the spiral arteries – a process critically dependent on decidual natural killer cells and decidual macrophages.^2–4^ By the late first trimester, maternal blood flows freely into the intervillous space, exposing the placenta to any pathogens or inflammatory mediators in maternal circulation.^5,6^ Despite this, the syncytiotrophoblasts lining the chorionic villi establish a barrier between maternal and fetal blood that largely protects the fetal compartment from direct microbial invasion. Placental maturation continues throughout the second and third trimesters through progressive branching of the villous tree, vascular remodeling, and establishment of a low-resistance, high-capacitance fetoplacental circulation required to support fetal growth.^1^ These developmental processes require sustained immune homeostasis at the maternal-fetal interface. Regulatory T cells and decidual macrophages remain essential in the second half of pregnancy, maintaining a microenvironment that supports immune tolerance to the semi-allogeneic fetus.^7–9^ Hofbauer cells – fetal-origin macrophages within the villous stroma – are essential to fetoplacental angiogenesis, villous remodeling, and barrier integrity.^10–12^ Disruptions of these processes underlie a spectrum of pregnancy complications including fetal growth restriction (FGR), small-for-gestational age (SGA) birth, and hypertensive disorders of pregnancy (HDP).^13–17^

A growing body of evidence demonstrates that maternal systemic infections – even those that do not directly infect the placenta or fetus – are associated with adverse placental outcomes. A retrospective study of patients with antepartum sepsis who recovered prior to delivery demonstrated that sepsis was associated with a nearly two-fold increased odds of placental dysfunction, particularly HDP which occurred in 24% of the sepsis cohort.^18^ Another retrospective review of patients hospitalized for a severe infection in the second half of pregnancy (primarily pyelonephritis and pneumonia) found increased rates of FGR, placental abruption, oligohydramnios, stillbirth, and perinatal mortality.^19^ Similarly, maternal SARS-CoV-2 infection, which rarely results in detectable placental viral invasion – has been linked with HDP particularly among nulliparous women^20–22^ and associated placental pathology including small placental weight and maternal-vascular malperfusion.^23^

Experimental models of maternal immune activation (MIA) support the biological plausibility that systemic maternal inflammation can alter placental immune and vascular pathways. Administration of low-dose lipopolysaccharide (LPS) to pregnant rats results in fetal growth restriction, deficient trophoblast invasion and spiral artery remodeling, altered uteroplacental hemodynamics, and features of preeclampsia including increased maternal blood pressure and proteinuria.^24^ Similarly, administration of viral mimetics such as PolyI:C to pregnant rodents induces upregulation of innate immune, interferon, cytokine, and antiviral response pathways in placental tissue, suggesting that the placenta itself mounts a substantial immune response to maternal systemic inflammation even in the absence of direct infection.^25^ These findings underscore a central paradox: a systemic infection that does not breach the placental barrier can nonetheless inflict significant placental damage, yet the cell-type-specific molecular mechanisms mediating this injury remain poorly understood.

Here, we use spatial transcriptomics to dissect cell-type-specific transcriptional changes in the human placenta following maternal pyelonephritis infection. Pyelonephritis is a bacterial infection in which microbes from the lower urinary tract ascend to the renal parenchyma. In some cases, pyelonephritis can result in maternal bacteremia, but the infection is not thought to directly invade placental tissue or the fetal compartment.^5^ By resolving the transcriptomes of distinct placental villous cell populations, we test whether systemic maternal infection, without direct placental invasion, reprograms the trophoblast, immune, and vascular compartments of the human placenta.

## Results

### Patient characteristics

A total of 14 placentas were included, from 6 pregnancies complicated by pyelonephritis and 8 control pregnancies. Among pyelonephritis cases, the mean (SD) gestational age at infection was 19.8 (8.8) weeks. Other maternal and pregnancy characteristics were similar between study groups (**Table 1**). All pregnancies delivered at term (mean [SD] gestational age at delivery, 38.9 [0.6] vs 38.8 [1.3] weeks in cases vs controls). There were no significant differences in rates of nulliparity (66.7% vs 62.5%, *P* = 1.00) or fetal sex distribution (33.3% vs 50.0% female, *P* = 0.63). Maternal comorbidities and delivery factors that could influence placental biology—including obesity, chronic hypertension, HDP, pregestational diabetes, gestational diabetes, labor prior to delivery, and clinical chorioamnionitis—were similar between groups.

**Table 1:** Comparison of clinical characteristics and obstetric outcomes.

| Variable | Pyelonephritis<br>(n=6) | Controls<br>(n=8) | p-value |
| --- | --- | --- | --- |
| Maternal age – years, mean (SD) | 27.3 (7.9) | 33.4 (6.7) | 0.18 |
| Nulliparous | 4 (66.7) | 5 (62.5) | 1.00 |
| Obesity (BMI ≥30) | 1 (16.7) | 0 (0.0) | 0.43 |
| Chronic hypertension | 0 (0.0) | 0 (0.0) | N/A |
| Pregestational diabetes | 0 (0.0) | 0 (0.0) | N/A |
| Female fetus | 2 (33.3) | 4 (50.0) | 0.63 |
| Gestational age at delivery – weeks, mean (SD) | 38.9 (0.6) | 38.8 (1.3) | 0.70 |
| Preterm birth (<37 weeks) | 0 (0.0) | 0 (0.0) | N/A |
| Hypertensive disorder of pregnancy | 1 (16.7) | 1 (12.5) | 1.00 |
| Gestational diabetes | 0 (0.0) | 0 (0.0) | N/A |
| Fetal growth restriction | 0 (0.0) | 0 (0.0) | N/A |
| Placental abruption | 1 (16.7) | 0 (0.0) | 0.43 |
| Clinical chorioamnionitis | 1 (16.7) | 0 (0.0) | 0.43 |
| Labor prior to delivery | 5 (83.3) | 6 (75.0) | 1.00 |
| Vaginal delivery | 2 (33.3) | 5 (62.5) | 0.57 |

Data are presented as n (%) for categorical variables and mean (SD) for continuous variables. P-values were calculated using the Fisher's exact test for categorical variables and the Wilcoxon rank-sum test for continuous variables. BMI, body mass index (kg/m<sup>2</sup>).

### Validation and deconvolution of morphology-defined spatial compartments

We performed GeoMx digital spatial profiling (DSP) of placental villi, using morphology-guided segmentation to profile PanCK-enriched trophoblast, CD31-enriched fetal endothelial cell, and CD68-enriched Hofbauer cell regions (**Figure 1a**). These segmented areas of illumination (AOIs) were analyzed separately to characterize transcriptional programs within each villous microenvironment.

**Figure 1.**
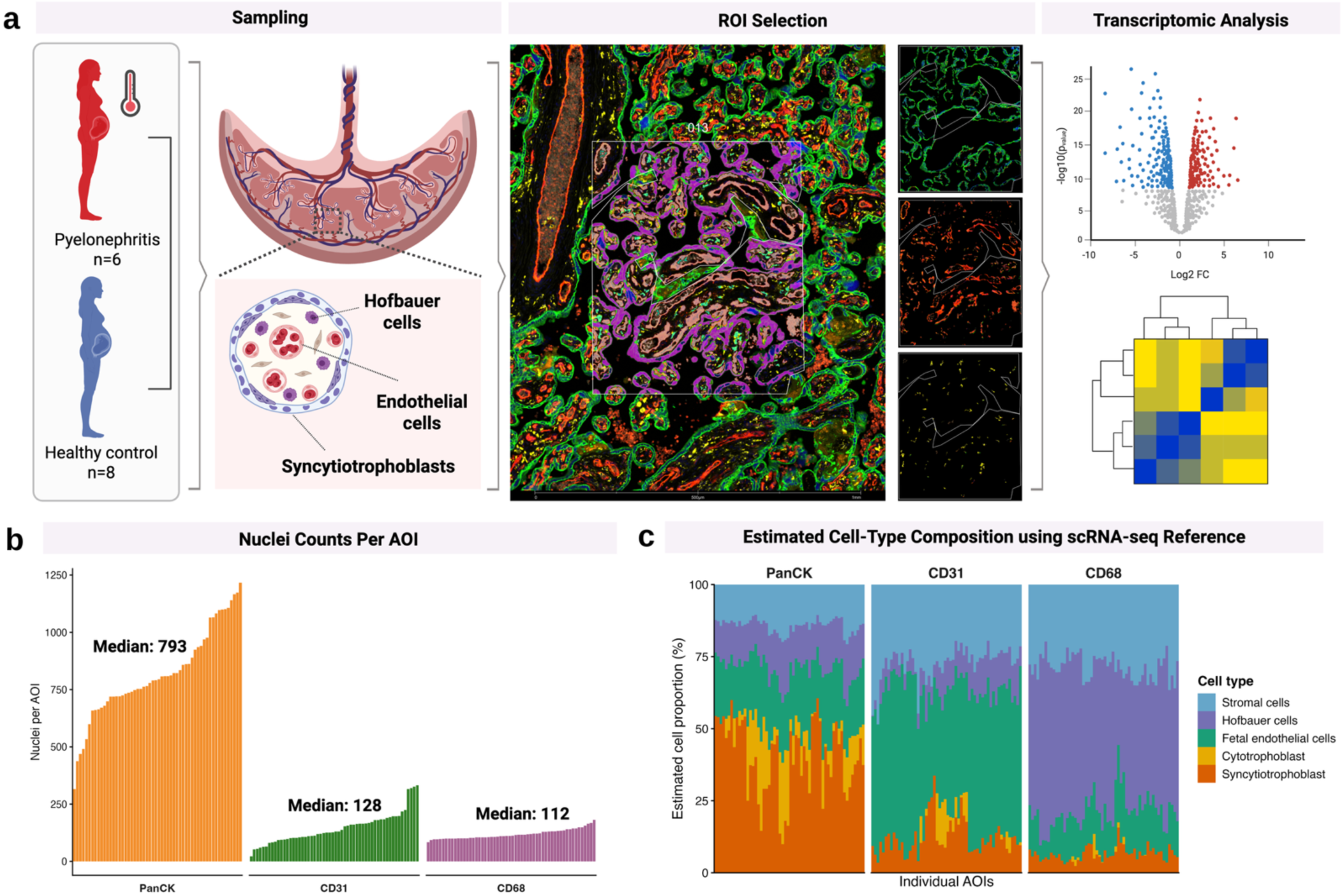
Study design and overview of spatial transcriptomic workflow. **a,** Experimental design. Placental samples from pregnancies complicated by antepartum pyelonephritis (n=6) and healthy controls (n=8) underwent GeoMx Digital Spatial Profiler whole-transcriptome profiling. Representative image shows segmentation of villous regions by morphology markers: trophoblast cells (PanCK, green), fetal endothelial cells (CD31, red), and Hofbauer cells (CD68, yellow). **b,** Nuclei counts per AOI across PanCK, CD31 and CD68 segments. **c,** SpatialDecon estimates of cell-type enrichment across morphology-defined segments using a human placental single-cell RNA-sequencing reference.

Following quality control (**Figure S1**), all 168 AOIs from 14 placentas were retained for downstream analyses. Probe- and gene-level filtering retained 17,797 genes for differential expression analysis. The mean area of each AOI was 35,530 μm^2^. The median nuclei counts per AOI were 793 for PanCK, 128 for CD31 and 112 for CD68 (**Figure 1b**). Morphology-defined segments showed clear transcriptional separation by principal component analysis (PCA) (**Figure 2a**) and enrichment of canonical lineage markers in their corresponding segments (**Figure 2b**). Pairwise differential expression demonstrated extensive transcriptional differences between segments (**Figure 2c**), while unsupervised clustering independently grouped AOIs predominantly by morphology-defined segment rather than study group (**Figure 2d**). Together, these findings indicate that cellular compartment was the dominant source of global transcriptional variation across placental AOIs.

**Figure 2.**
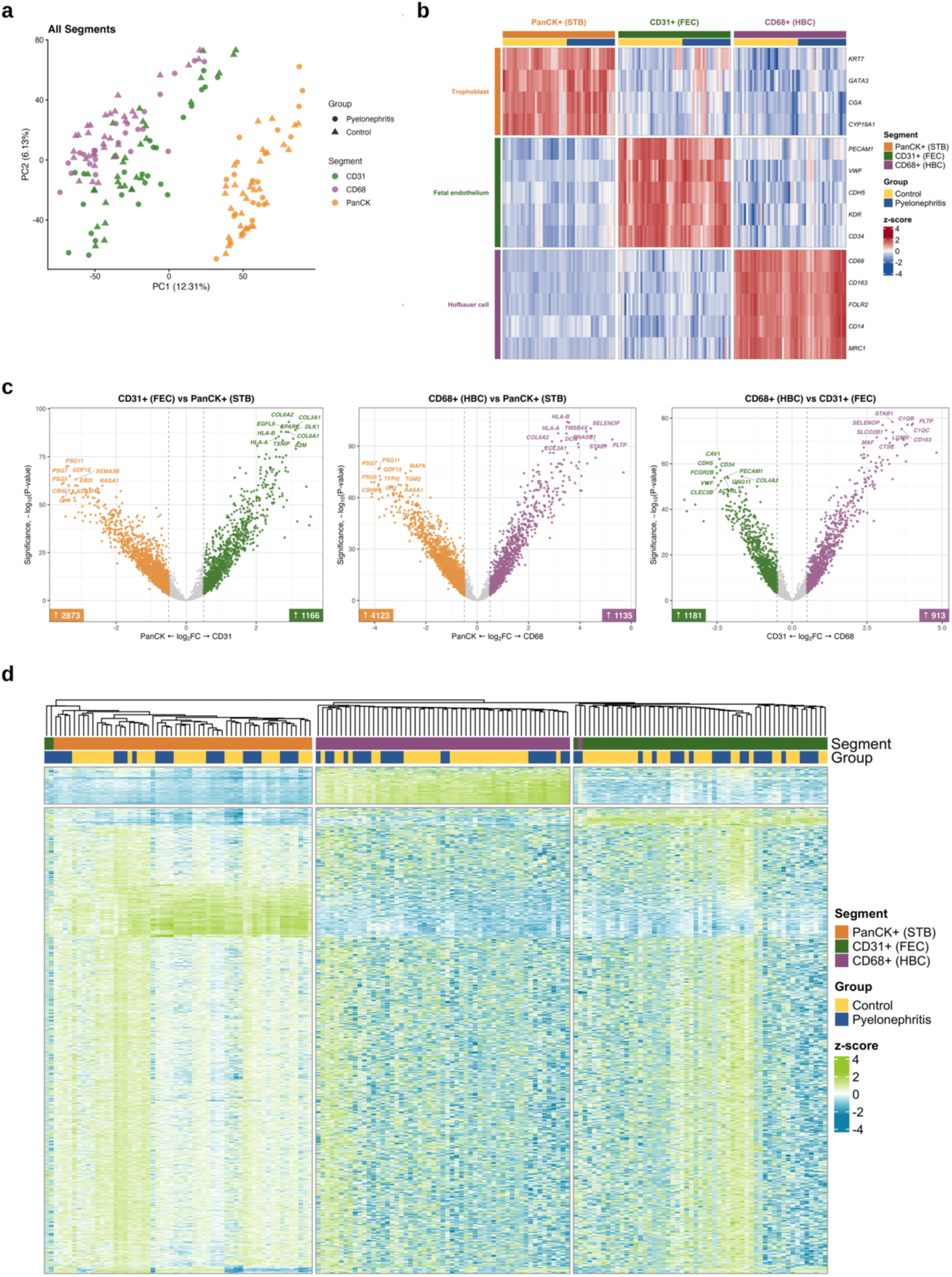
Morphology-guided segmentation yields transcriptionally distinct placental compartments. **a,** Principal component analysis (PCA) of log2 Q3-normalized expression across 168 AOIs (56 per segment; 14 placentas), colored by morphology-defined segment and shaped by study group. **b,** Canonical trophoblast, fetal endothelial, and Hofbauer cell markers were enriched in their corresponding morphology-defined segments, supporting appropriate lineage enrichment. Values represent row-scaled log2 Q3-normalized expression. **c,** Pairwise differential expression demonstrated extensive transcriptional differences between morphology-defined segments. Genes meeting FDR < 0.05 and |log2FC| > 0.5 are colored by the segment in which expression was higher; numbers indicate significant genes in each direction. **d,** Unsupervised hierarchical clustering of the 10% most variable genes predominantly grouped AOIs by morphology-defined segment rather than study group, further demonstrating that cellular compartment was the dominant source of global transcriptional variation. Columns are annotated by segment and study group.

Reference-based spatial deconvolution further confirmed enrichment of the expected dominant cell populations within each morphology-defined segment (**Figure 1c**). PanCK AOIs were enriched for syncytiotrophoblasts and cytotrophoblasts, CD31 AOIs for fetal endothelial cells, and CD68 AOIs for Hofbauer cells, while each compartment also contained contributions from neighboring villous cell populations. This pattern is consistent with the known architecture of placental villi, in which fetal blood vessels, stromal fibroblasts, Hofbauer cells, and cytotrophoblasts are organized in close spatial proximity within the villous core. GeoMx DSP uses fluorescence-based morphology markers to define cell-enriched segments within each tissue section; however, because transcript capture occurs across the full thickness of the section, transcripts from closely apposed cells along the z-axis may contribute to the same AOI. Rather than representing isolated cell populations, the segmented AOIs therefore capture physiologic villous niches, representing trophoblast, fetal endothelial cell, and fetal Hofbauer cell microenvironments.

Additionally, reference-based spatial deconvolution infers cell identity from transcriptional similarity, therefore overlapping gene-expression programs between placental cell populations may contribute to mixed cellular assignments. For instance, trophoblasts expressing vascular-associated transcriptional programs may be assigned to endothelial cell populations within the reference atlas.^4,26–28^ The spatial deconvolution results should be interpreted as estimates of the cellular composition of each AOI rather than definitive assignments of individual cell identities.

There were no significant differences in cell-type composition between infection and control samples (**Figure S2**), supporting that infection-associated transcriptional differences likely reflect alterations in cellular activation states or intercellular signaling rather than shifts in cellular abundance.

### Pyelonephritis induces compartment-specific transcriptional responses

Differential expression analysis revealed marked heterogeneity in the placental response to pyelonephritis across morphology-defined compartments. Using an exploratory threshold of unadjusted p<0.05 and |log_2_FC| ≥ 0.5, 419 genes were identified in CD68-segmented AOIs, compared with 206 in CD31 and 23 in PanCK segments (**Figure 3b**). Overlap between compartments was limited, with only three differentially expressed genes (DEGs) shared across all three compartments (**Figure 3c,d**), indicating that pyelonephritis elicits predominantly compartment-specific transcriptional responses within placental villi.

**Figure 3.**
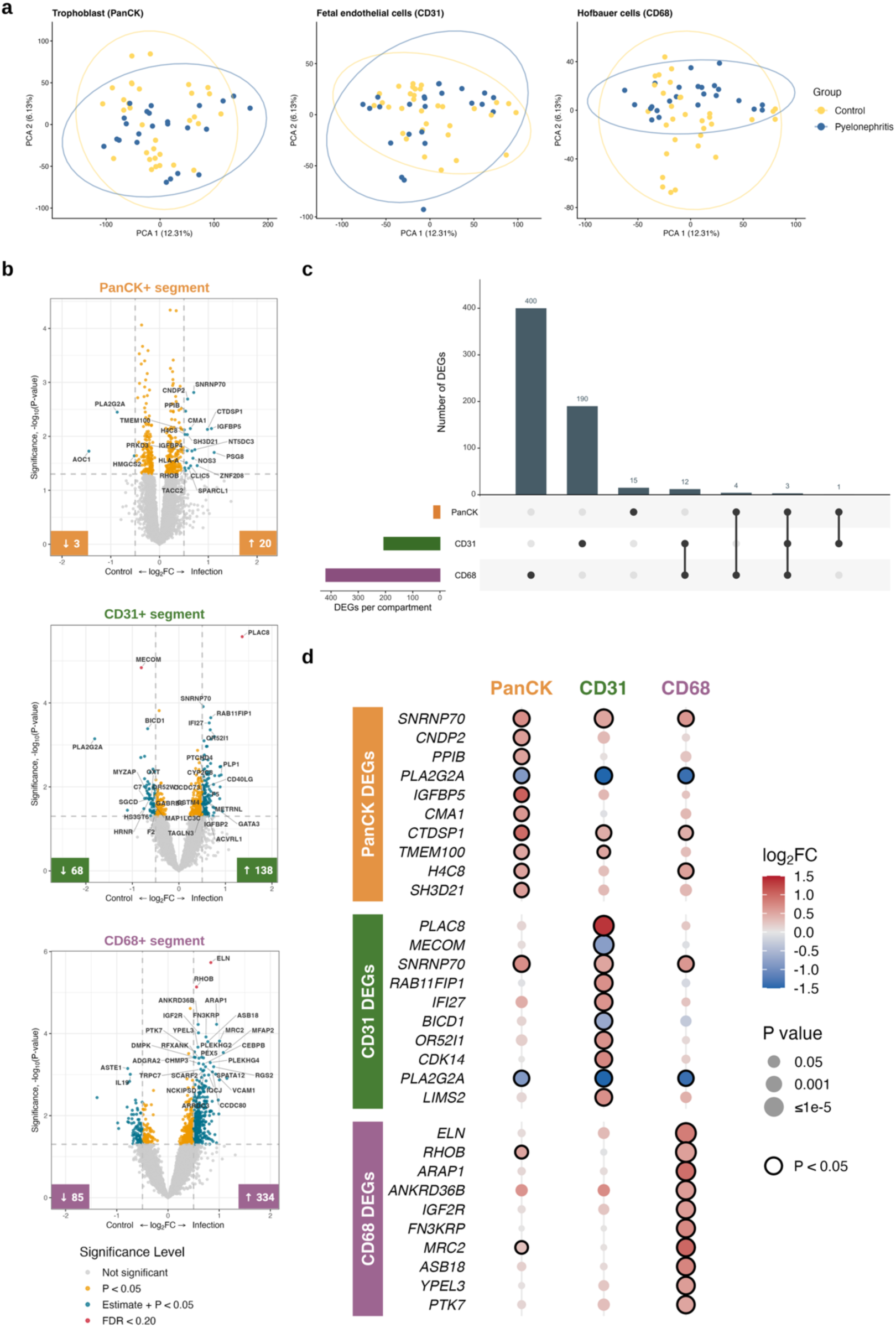
Pyelonephritis induces compartment-specific placental transcriptional responses. **a,** PCA of each morphology-defined segment, colored by study group; groups overlapped substantially within each segment with partial separation in the CD68+ Hofbauer-cell enriched AOIs. Ellipses indicate the 95% normal-distribution regions. **b,** Volcano plots comparing pyelonephritis and control placentas within each segment. DEGs were defined as *P* < 0.05 and |log2FC| ≥ 0.5; numbers indicate downregulated (↓) and upregulated (↑) genes meeting these thresholds. Dot colors indicate significance. **c,** UpSet plot showing limited overlap of DEGs among segments, with only three genes shared across all three segments. **d,** Top 10 DEGs in each segment, ranked by *P* value, with differential expression results shown across all three segments.

### Hofbauer cell-enriched segments demonstrate coordinated extracellular matrix remodeling programs

The CD68-positive Hofbauer cell-enriched compartment exhibited the most extensive transcriptional response to antepartum pyelonephritis (**Figure 3a,b**). Among the individual DEGs, the *ELN* gene (encoding elastin) represented a particularly prominent finding and was significantly increased in infection-associated placentas (log_2_FC = 0.84, FDR = 0.033). *ELN* also contributed to multiple enriched biological processes, indicating that elastin upregulation formed part of a coordinated extracellular matrix (ECM) remodeling network rather than an isolated gene-level observation. Gene set enrichment analysis (GSEA) demonstrated coordinated activation of extracellular matrix organization (**Figure 4a**) (Normalized Enrichment Score [NES] = 1.72), collagen metabolism (NES = 1.72), cell-substrate adhesion (NES = 1.62), wound healing (NES = 1.41), and TGF-β receptor signaling (NES = 1.49). Ingenuity Pathway Analysis (IPA) similarly identified a coordinated matrix-remodeling program (**Figure 4c**), with predicted activation of extracellular matrix organization (z = 3.00), elastic fiber formation (z = 2.65), integrin cell-surface interactions (z = 2.45), wound healing (z = 2.11), and pathways involved in collagen assembly, biosynthesis, and degradation (z = 2.00 for each).

**Figure 4.**
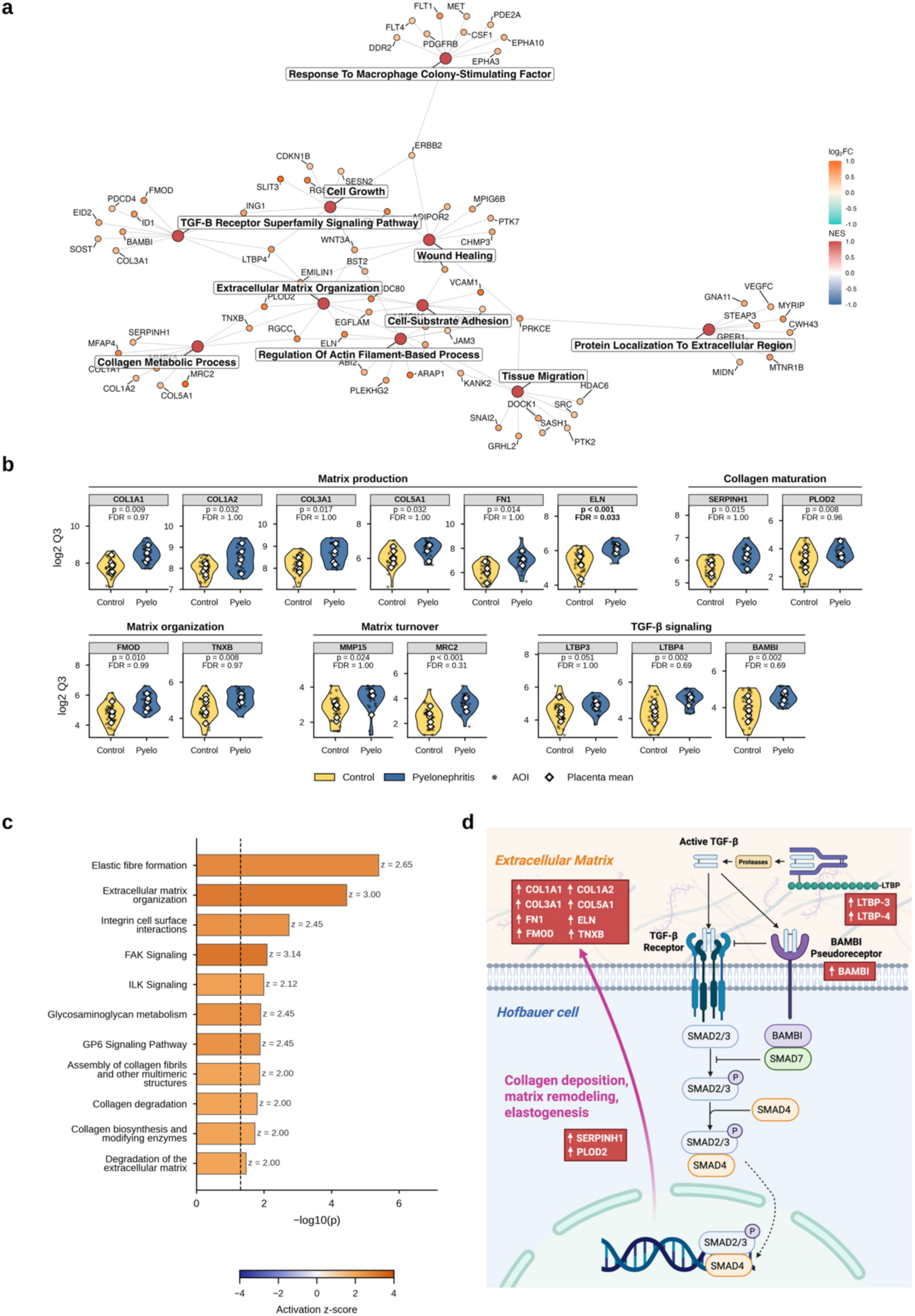
Pyelonephritis is associated with coordinated extracellular matrix and TGF-β remodeling programs in Hofbauer cell-enriched regions. **a,** Gene–concept network (cnetplot) of selected GSEA pathways enriched in CD68+ AOIs. Pathway nodes are colored by NES and gene nodes by log_2_FC. **b,** Expression of selected matrix- and TGF-β-related genes. Points represent AOIs and diamonds placenta means. **c,** Selected extracellular matrix-related IPA pathways. Bars indicate −log10 *P* value and colors indicate activation z-scores. **d,** Schematic of TGF-β/SMAD signaling and its regulation by genes altered in Hofbauer cell-enriched regions. Canonical TGF-β signaling activates SMAD2/3– SMAD4-mediated transcription, with *LTBP3/4* regulating extracellular TGF-β availability and *BAMBI* negatively regulating receptor signaling. Genes increased in pyelonephritis are shown in red. *Created with BioRender*.

Analysis of leading-edge genes contributing to the enriched GSEA pathways demonstrated increased expression of genes involved in extracellular matrix production and deposition (**Figure 4b**), including structural collagen genes (*COL1A1, COL1A2, COL3A1, COL5A1*), together with genes encoding elastic fiber assembly *(ELN, EMILIN1, MFAP2)* and matrix scaffolding *(FN1, FMOD, VCAN).* This ECM synthesis program was accompanied by increased expression of matrix maturation genes *(SERPINH1, PLOD2)*. In parallel, genes regulating matrix turnover were enriched including the matrix metalloproteinase *MMP15* and the collagen endocytic receptor *MRC2*, indicating that matrix remodeling occurred alongside new matrix deposition. Collectively, our findings demonstrate a coordinated extracellular matrix remodeling program characterized by increased matrix production, maturation and turnover following antepartum pyelonephritis.

The remodeling signature also converged on altered regulation of TGF-β signaling (**Figure 4d**). Increased expression of *LTBP3* and *LTBP4*, which regulate extracellular sequestration and activation of latent TGF-β complexes, suggests that Hofbauer cells actively modulate local TGF-β bioavailability. This occurred alongside increased expression of *BAMBI*, a transmembrane pseudoreceptor whose extracellular domain resembles TGF-β type I receptors but whose intracellular domain lacks the serine/threonine kinase domain required for signal transduction. *BAMBI* is directly induced by TGF-β signaling through a SMAD3-SMAD4-dependent transcriptional mechanism and subsequently inhibits signaling by blocking functional receptor complex formation and cooperating with SMAD7 to prevent R-SMAD phosphorylation, thereby providing negative feedback on further TGF-β pathway activation. Collectively, these findings demonstrate coordinated transcriptional regulation of the TGF-β signaling network, including extracellular ligand availability and pathway-associated regulatory mechanisms.

### Histologic assessment identifies alterations in extracellular matrix organization in infection-associated placentas

To assess the tissue-level consequences of our transcriptomic findings, we performed Verhoeff-Van Gieson (VVG) staining to characterize elastic fiber and collagen organization in the perivascular contractile sheath (PVCS) and perivascular stroma (PVS) of fetal villous vessels (**Figure 5a)**. Vessel geometry did not differ between groups, including PVCS and PVS area normalized to lumen area (**Figure S3)**. Pyelonephritis-exposed placentas instead showed altered perivascular composition. Nuclear area fraction was higher in pyelonephritis placentas (2.82% vs 2.11%, *P* = 0.023), with consistent increases in both arteries (*P* = 0.036) and veins (*P* = 0.031) (**Figure 5b**). Elastin fraction showed a non-significant trend toward being higher (42% vs 39%, *P* = 0.095), and collagen fraction a trend toward being lower (52% vs 56%, *P* = 0.068) in pyelonephritis placentas. Among the 14 GeoMx participants, *ELN* expression in CD68+ Hofbauer cell-enriched AOIs correlated with perivascular elastin content (ρ = 0.67, *P* = 0.008) (**Figure 5c**). This association was strongest in the PVS (ρ = 0.76) and was consistent across arteries, veins, and stem villous vessels (ρ = 0.57–0.81) (**Figure 5d**).

**Figure 5.**
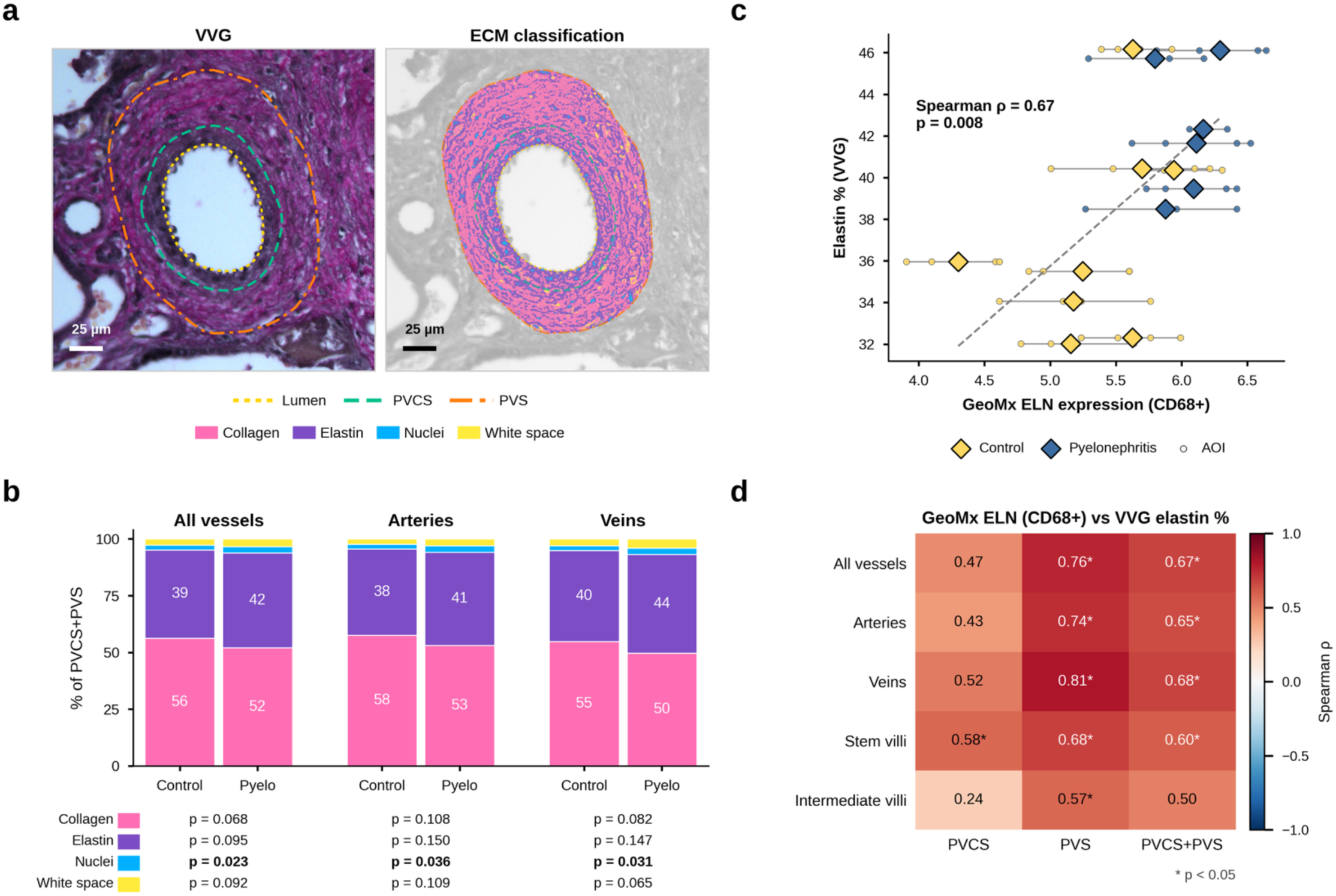
Perivascular extracellular matrix composition in villous vessels and its association with Hofbauer cell *ELN* expression. **a,** Representative VVG-stained vessel and corresponding pixel classification of the perivascular contractile sheath (PVCS) and perivascular stroma (PVS) into collagen, elastin, nuclei, and white space. **b,** Mean composition of the combined perivascular zone (PVCS+PVS), stratified by study group and vessel type. Pyelonephritis was associated with significantly increased nuclear content across vessel types and with a non-significant shift toward greater elastin and lower collagen. *P* values are from linear mixed-effects models. **c,** Association between *ELN* expression from CD68+ GeoMx AOIs and VVG elastin fraction in the combined perivascular zone (Spearman ρ = 0.67, *P* = 0.008). GeoMx expression values are log2 Q3-normalized counts. Circles represent individual GeoMx AOIs and diamonds placenta means. **d,** Correlations between *ELN* expression from CD68+ GeoMx AOIs and VVG elastin fraction across perivascular regions, vessel types, and villous levels. Values are Spearman ρ; asterisks indicate *P* < 0.05.

### Antepartum pyelonephritis alters placental vascular remodeling programs

The CD31-enriched (fetal endothelial) compartment demonstrated transcriptional alterations involving pathways relevant to vascular remodeling and function. At the individual-gene level, *PLAC8* represented the strongest individual finding and was significantly increased in infection-associated placentas (log_2_FC = 1.36, FDR = 0.047) (**Figure 3b)**. *PLAC8* is a hypoxia-responsive gene transcriptionally driven by HIF-1α, whose increased expression has been associated with impaired endothelial differentiation and angiogenic function through downregulation of NOX4-dependent signalling.^29,30^ *MECOM*, a transcriptional regulator of endothelial cell identity, was decreased (log_2_FC = −0.81, FDR = 0.129), which may indicate altered endothelial differentiation in infection-associated placentas.

Although no pathways met significance criteria by GSEA in the CD31-enriched compartment, IPA predicted activation of the RHO GTPase cycle (z = 2.83), a central regulator of actin cytoskeletal remodeling, cell-matrix adhesion, endothelial migration, and vascular morphogenesis (**Figure 6b**). In contrast, VDR/RXR activation was predicted to be inhibited (z = −2.00). VDR/RXR signaling supports endothelial homeostasis through anti-inflammatory, antioxidant, and vasoregulatory mechanisms, including regulation of endothelial nitric oxide signaling. PPAR signaling, a related RXR-dependent nuclear receptor pathway involved in metabolic homeostasis and vascular function, was also significantly enriched (*P* = 0.009) with a negative directional trend (z = −1.00), although it did not meet the threshold for predicted inhibition. Together, these findings suggest a shift toward a vascular remodeling phenotype, accompanied by attenuation of endothelial homeostasis programs.

**Figure 6.**
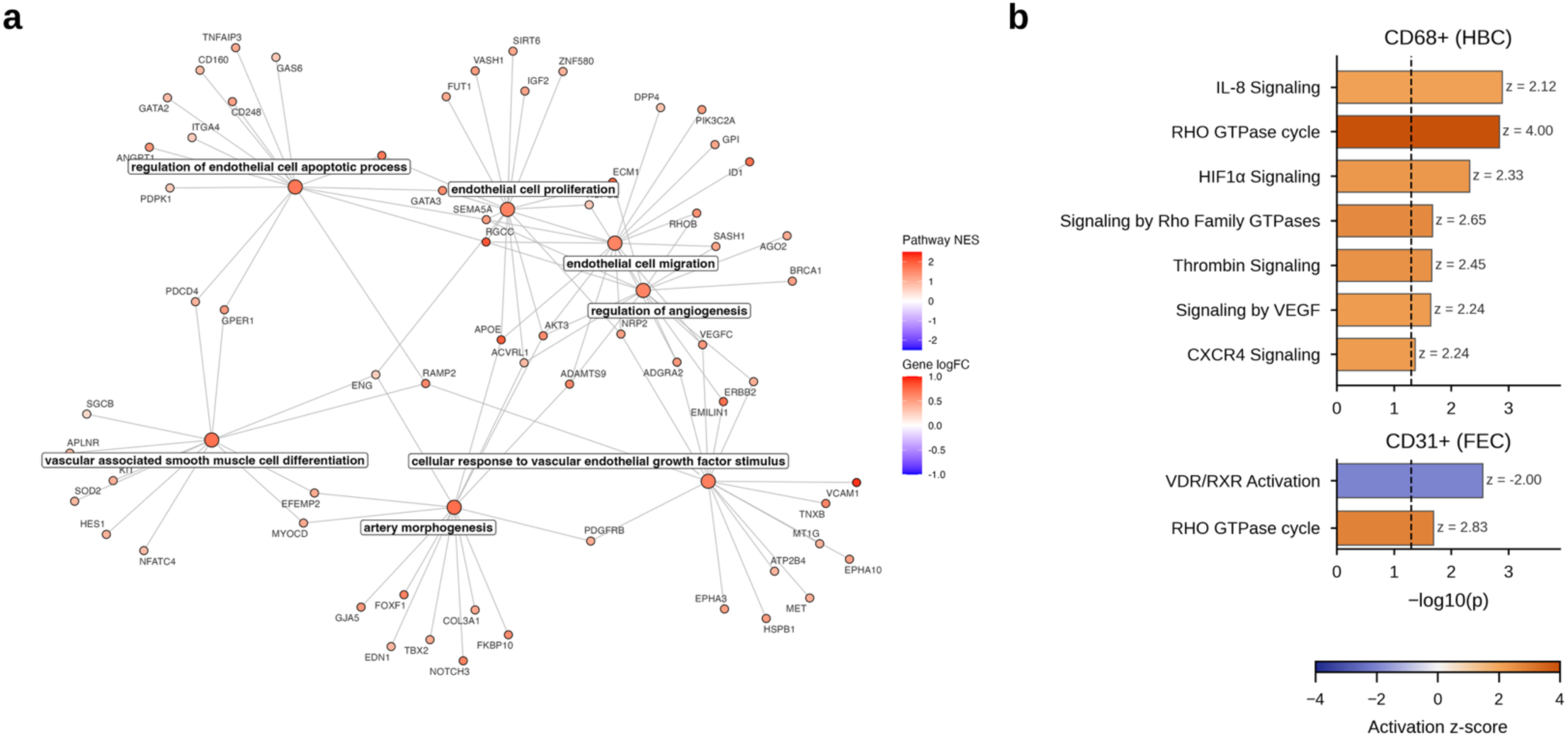
Pyelonephritis is associated with altered placental vascular signaling pathways. **a,** Gene–concept network (cnetplot) of selected GSEA vascular remodeling and endothelial signaling pathways enriched in CD68+ Hofbauer cell-enriched AOIs. Pathway nodes are colored by NES and gene nodes by log_2_FC. **b,** Selected vascular regulatory pathways identified by IPA in CD68+ Hofbauer cell-enriched (top) and CD31+ fetal endothelial-enriched (bottom) AOIs. Pathways shown meet *P* < 0.05 and |z-score| ≥ 2; bars indicate −log10 *P* value and color indicates activation z-score.

We next examined whether vascular regulatory programs extended beyond the fetal endothelial compartment. CD68-enriched (Hofbauer cell) segments demonstrated broad enrichment of vascular developmental programs (**Figure 6a,b)**. GSEA identified the following significantly enriched pathways: regulation of angiogenesis (NES = 1.58), positive regulation of endothelial cell migration (NES = 1.73), and regulation of endothelial cell apoptotic process (NES = 1.74). These pathways included leading-edge genes involved in angiogenic signaling and vascular development, including *VEGFC, RHOB, RGCC, ACVRL1, NRP2, AKT3*, and *EDN1*. IPA similarly predicted activation of VEGF signaling (z = 2.24) and HIF1α signaling (z = 2.33), consistent with a hypoxia-responsive program that paralleled *PLAC8* upregulation in fetal endothelial-enriched regions.

Histopathologic findings provided complementary tissue-level evidence of altered placental vascular remodeling in infection-associated placentas **(Table 2)**. Accelerated villous maturation, distal villous hypoplasia, and increased syncytial knotting—features associated with maternal vascular malperfusion (MVM) —were each more frequent in pyelonephritis-exposed placentas than in controls (38% vs 0%, *P* = 0.032 for each). Perivillous fibrin deposition, a marker of impaired intervillous perfusion and trophoblast injury, was also significantly increased (50% vs 0%, *P* = 0.008). In contrast, fetal vascular malperfusion (FVM) did not differ significantly between groups (25% vs 6.7%, *P* = 0.27), with all observed FVM lesions being low-grade and no fetal vascular thrombosis or vessel obliteration detected in any placenta. Histologic chorioamnionitis was also more frequent in pyelonephritis-exposed placentas (62.5% vs 13.3%, *P* = 0.03) despite similar rates of clinical chorioamnionitis between groups (25.0% vs 20.0%, *P* = 1.00; **Table S3**), suggesting subclinical acute inflammation of the fetal membranes.

**Table 2:** Comparison of histopathologic placental findings.

| **Variable** | **Pyelonephritis**<br>**(n=8)** | **Controls**<br>**(n=15)** | ***P*-value** |
| --- | --- | --- | --- |
| Maternal vascular malperfusion | 2 (25.0) | 0 (0) | 0.11 |
| Placental infarct | 0 (0) | 0 (0) | 1.00 |
| Accelerated villous maturation | 3 (37.5) | 0 (0) | **0.03** |
| Retroplacental hemorrhage | 1 (12.5) | 0 (0) | 0.35 |
| Distal villous hypoplasia | 3 (37.5) | 0 (0) | **0.03** |
| Syncytial knots | 3 (37.5) | 0 (0) | **0.03** |
| Fetal vascular malperfusion | 2 (25.0) | 1 (6.7) | 0.27 |
| Thrombosis | 0 (0) | 0 (0) | 1.00 |
| Avascular villi | 2 (25.0) | 0 (0) | 0.11 |
| Stromal-vascular karyorrhexis | 1 (12.5) | 0 (0) | 0.35 |
| Vessel obliteration | 0 (0) | 0 (0) | 1.00 |
| Vascular ectasia | 0 (0) | 0 (0) | 1.00 |
| Perivillous fibrin deposition | 4 (50.0) | 0 (0) | **0.008** |
| Chronic villitis | 0 (0) | 0 (0) | 1.00 |
| Deciduitis | 1 (12.5) | 0 (0) | 0.35 |
| Histopathologic chorioamnionitis | 5 (62.5) | 2 (13.3) | **0.03** |
| Meconium staining | 2 (25.0) | 2 (13.3) | 0.59 |

Placental histopathologic findings were characterized according to the Amsterdam consensus criteria. Data are presented as n (%), unless otherwise specified. *P*-values were calculated using the Fisher exact tests.

### Trophoblast transcriptional profiles remain largely unchanged following pyelonephritis

The PanCK+ trophoblast-enriched compartment showed minimal transcriptional response to antepartum pyelonephritis (**Figure 3**). No gene reached statistical significance after correction for multiple testing (minimum FDR = 0.42 across 17,797 genes), and only 376 genes (2.1%) reached nominal significance (*P* < 0.05), fewer than expected by chance alone. Consistent with the limited gene-level signal, GSEA identified no significantly enriched biological processes, while IPA identified only overlap-based canonical pathway associations driven by two to three genes and without activation z-scores.

## Discussion

Maternal pyelonephritis was associated with cell-type specific alterations in the placental transcriptome. The most extensive response occurred within Hofbauer cell-enriched regions, where coordinated programs of ECM organization, matrix turnover, TGF-β regulation and vascular development were enriched. Fetal endothelial-enriched regions showed more limited changes involving endothelial identity and vascular remodeling, whereas the sampled trophoblast-enriched regions demonstrated little detectable transcriptional response. Notably, these differences were detected in placentas collected at delivery, after clinical recovery from the antepartum infection, suggesting that resolution of maternal illness may not coincide with normalization of placental transcriptional programs. These findings provide molecular context for the association between maternal systemic infection and placental dysfunction,^18^ implicating the villous immune–stromal and vascular microenvironments as sites of infection-associated alterations.

The dominant transcriptomic finding was a coordinated ECM remodeling signature within Hofbauer cell-enriched niches. Increased expression of structural matrix genes occurred alongside genes involved in matrix assembly, maturation and degradation, suggesting a response encompassing multiple stages of matrix remodeling. *ELN* – encoding elastin – emerged as a consistent signal across two independent experimental approaches: its upregulation in Hofbauer cell-enriched regions remained significant after multiple-testing correction, and GeoMx *ELN* expression correlated with perivascular elastin content measured by blinded VVG analysis of independently sampled vessels from the same placentas. An expanded VVG cohort incorporating additional pyelonephritis and control placentas beyond the GeoMx cohort showed a similar trend toward higher elastin content in the perivascular regions of infection-associated placentas. Together, these findings support elastin-associated remodeling at both the transcriptional and tissue levels.

The ECM signature within Hofbauer-cell enriched regions could reflect direct matrix production by macrophages. In human atheroma and abdominal aortic aneurysms, macrophages synthesize ECM components including tropoelastin, but the matrix they produce tends to be disorganized compared to that produced by smooth muscle cells.^31^ Alternatively, our transcriptomic findings may reflect macrophage–fibroblast interactions, with matrix production by adjacent fibroblasts. In pulmonary hypertension, cardiac fibrosis and atherosclerosis, reciprocal signaling between these cell populations regulates inflammatory states, fibroblast activation and ECM remodeling.^32–39^ Macrophages can influence fibroblasts through TGF-β, PDGF and inflammatory cytokines, while fibroblasts can shape macrophage behavior through secreted mediators and changes in the surrounding matrix.^40–44^ Matrix stiffness and proteolytic fragments can further modify cellular signaling, creating feedback between immune activation and tissue remodeling. The TGF-β-associated patterns observed in our transcriptomic data are consistent with this framework. *LTBP3* and *LTBP4* participate in the secretion and extracellular localization of latent TGF-β, potentially modifying the pool available for subsequent activation.^45^ *LTBP4* additionally promotes incorporation of elastin into the microfibrillar scaffold, linking latent growth-factor regulation to elastic fiber assembly.^46^ In contrast, *BAMBI* inhibits TGF-β receptor signaling and can itself be induced by TGF-β, making its increased expression consistent with feedback that limits further pathway responsiveness.^47,48^

Whether the observed ECM remodeling is adaptive or maladaptive remains unresolved. Maternal vascular malperfusion and perivillous fibrin deposition which we observed histologically could impair oxygen and nutrient exchange, creating an impetus for compensatory remodeling within the villous core. Matrix turnover and vascular growth programs might help accommodate these disturbances and preserve exchange capacity. Alternatively, excessive or disorganized matrix remodeling could alter tissue mechanics and impair vascular development or function. Our findings may therefore reflect residual injury or a reparative response.

A potential upstream contributor to these changes lies on the maternal side of the interface. Pyelonephritis-exposed placentas showed more intervillous macrophage aggregates adjacent to sampled villi (29.2% vs 6.3% of ROIs; **Figure S4b**). Because ROIs were restricted to villous tissue, these cells were excluded from our transcriptomic profiles and their origin was not directly assessed. However, macrophages within the intervillous space are generally of maternal origin, and their accumulation may reflect persistent maternal immune activation at the maternal-fetal interface after clinical resolution of infection.^6^ Intervillous immune cell infiltration has been associated with disruption of the intervillous space and adverse pregnancy outcomes and could therefore contribute to the villous stromal and vascular remodeling observed.^49–51^

Our Hofbauer cell- and fetal endothelial-enriched compartments also demonstrated transcriptional changes related to vascular development and remodeling. Within Hofbauer cell-enriched regions, enriched pathways included endothelial migration, VEGF signaling, HIF-1α signaling, and other programs involved in angiogenic regulation. The CD31-enriched compartment showed changes in genes involved in endothelial identity and vascular remodeling, including coordinate *PLAC8* upregulation and *MECOM* downregulation, raising the possibility of altered endothelial identity and a response to hypoxic stress. These findings may be mechanistically connected. The ECM provides the structural and biochemical scaffold within which placental vessels develop, regulating endothelial adhesion, migration, and angiogenesis.^52,53^ Prior studies demonstrate that ECM derived from FGR placentas can impair endothelial migration and angiogenesis, whereas healthy placental ECM can restore these functions.^54,55^ Thus, the prominent ECM remodeling observed within Hofbauer cell-enriched niches could potentially alter the microenvironment in which adjacent fetal endothelial cells develop and function.

The limited transcriptional response in trophoblast-enriched regions may have several explanations. Trophoblast transcriptional programs may be less affected by antecedent maternal infection than those of other villous compartments. This finding is consistent with prior placental transcriptomic studies, in which the signal attributable to antecedent maternal infection is concentrated in maternal immune and fetal stromal compartments rather than trophoblast populations.^56^ Tissue selection may also have contributed. Areas with substantial fibrin deposition were deliberately avoided to obtain adequate transcript signal (**Figure S4)**, potentially biasing the sampled regions toward relatively healthy trophoblast cells, particularly given the substantial frequency of perivillous fibrin deposition in infection-associated placentas (**Table 2**). Thus, the limited differential expression observed in trophoblast-enriched regions does not exclude broader trophoblast alterations across the placenta.

### Strengths and Limitations

Strengths of this study include spatially resolved profiling of human placental tissue, enabling comparison of trophoblast-, Hofbauer cell-, and fetal endothelial-enriched villous niches while preserving their tissue context. PCA and reference-based deconvolution supported enrichment of the intended cellular compartments, and the absence of significant differences in estimated cell-type proportions between groups reduced the likelihood that the major transcriptional findings were driven by differences in cellular composition. Clinical characteristics and baseline medical comorbidities were broadly comparable between groups, reducing the risk of confounding by maternal conditions associated with both infection and placental dysfunction. A further strength was the convergence of independent approaches on ECM remodeling: *ELN* remained significant after multiple-testing correction, ECM-related programs were identified across pathway analyses, and increased *ELN* expression was corroborated at the tissue level by VVG analysis.

Several limitations should be considered. The small number of independent placentas limited statistical power; multiple AOIs increased spatial sampling but did not increase the number of independent biological samples. Accordingly, findings require validation in larger cohorts. Cases also varied in gestational age at infection and transcriptomic profiling occurred only at delivery, limiting assessment of the temporal evolution of these changes. The observational design further precludes causal attribution of the placental findings to pyelonephritis.

Tissue selection remains an important consideration, as some pathologic placental regions are inherently less amenable to spatial transcriptomic profiling. Although the distribution of ROIs adjacent to fibrin did not differ between groups (**Figure S4b)**, fibrin deposition was more prevalent overall in pyelonephritis placentas (**Table 2)**. Specific pathologic lesions, including avascular villi, fibrin deposition, and distal villous hypoplasia, may contain insufficient cellular material for reliable transcriptomic analysis and are therefore inherently underrepresented. Macrophage-enriched regions also required sufficient nuclei for profiling, potentially favoring macrophage-dense villous niches. Finally, although ROIs contained mixtures of villous types and the predominant villous type did not differ significantly between groups **(Figure S4b)**, ECM composition varies across the placental vascular tree, and differences in the representation of stem versus intermediate and terminal villi could have influenced the observed matrix signatures.

Finally, GeoMx segments were cell-enriched rather than cell-specific, and transcripts from neighboring cells could contribute to each AOI. Orthogonal validation approaches such as multiplex RNAscope or single-cell RNA sequencing could confirm the cellular sources of key signals. The study also did not directly measure TGF-β activity, elastic fiber organization, endothelial function, or vascular reactivity; thus, the functional and mechanistic implications of the observed transcriptional programs remain hypothesis-generating.

In summary, spatially resolved profiling of the term human placenta reveals that antecedent maternal pyelonephritis leaves a durable, compartment-specific imprint on villous biology that persists after clinical recovery. Rather than a uniform placental response, pyelonephritis was associated predominantly with transcriptional alterations in Hofbauer cell-enriched and fetal vascular niches, with limited changes in trophoblast-enriched regions. The convergence of extracellular matrix, TGF-β and vascular developmental programs with independent evidence of altered perivascular matrix composition on VVG staining implicates immune–stromal–vascular interactions as a potential organizing axis of this response. Our findings carry broader implications for obstetric management: normalization of maternal clinical status may not reflect normalization of placental biology. Whether these changes represent residual injury or compensatory adaptation remains to be defined. Larger cohorts and experimental validation of the mechanism linking immune-stromal signaling to matrix remodeling and vascular function will be needed to establish the reproducibility and biological significance of our findings.

## Material and Methods

### Study Design

We performed a retrospective case-control analysis of human placental formalin-fixed paraffin-embedded (FFPE) tissue to investigate transcriptional alterations associated with severe antepartum pyelonephritis infection. Cases and controls were defined a priori based on clinical phenotype and were selected from the same source population. A total of 14 placental samples were included, comprising 6 cases and 8 controls.

Spatial transcriptomic profiling was performed using the GeoMx® Digital Spatial Profiler (Bruker Corporation). Regions of interest (ROIs) were defined to include only chorionic villi, with intervillous spaces excluded. Immunofluorescent morphology markers were used to define cell-type-enriched areas of illumination (AOIs), including *PanCK* (trophoblast), *CD68* (macrophage), and *CD31* (endothelial cell) compartments. The primary exposure was antepartum pyelonephritis, and the primary outcome was gene expression within each cell-type-enriched compartment. Investigators were blinded to clinical group during ROI and AOI selection. The study was observational in nature and not designed to establish causality.

### Study Population and Tissue Processing

Placental samples were obtained from participants receiving clinical care at the University of California, San Francisco (UCSF) and San Francisco General Hospital. All participants provided written informed consent, under protocols approved by UCSF Institutional Review Board (#21-33576; #20-32077; #20-30410). Specimens were collected between June 2020 and May 2024. Inclusion criteria included delivery of a liveborn fetus at ≥28 weeks’ gestation. Pregnancies complicated by major congenital or genetic anomalies were excluded.

Cases were defined as participants hospitalized for treatment of suspected pyelonephritis with documented bacteriuria consistent with a urinary tract infection. All cases demonstrated clinical recovery prior to delivery, defined as discharge from the antepartum infection hospitalization prior to delivery. Controls were defined as participants without an antepartum infection requiring an emergency department visit or hospitalization. Mild infections managed in the outpatient setting, including asymptomatic bacteriuria or uncomplicated urinary tract infections, did not preclude inclusion as controls. Participant demographic and clinical variables were abstracted from the electronic health record using prespecified definitions (**Table S1**) by investigators blinded to downstream molecular readouts. Placentas were collected within 3 hours after delivery, fixed in neutral-buffered formalin for 24–32 hours, washed in phosphate-buffered saline (PBS), and stored in 70% ethanol at 4°C prior to paraffin embedding (FFPE).

### Transcriptomic profiling experiments

Transcriptomic profiling was performed using the GeoMx Digital Spatial Profiler, according to the manufacturer’s recommended protocol, as previously described.^57^ One FFPE tissue block was obtained from each participant. Biological replicates were defined at the participant level, with ROIs and AOIs treated as repeated measures nested within individuals.

Slides containing 5μm-thick FFPE sections were deparaffinized, rehydrated, and subjected to antigen retrieval in Tris-EDTA reagent (99°C), followed by proteinase K treatment (37 °C) to expose RNA targets. After washing, slides were hybridized overnight with oligo-tagged in situ hybridization probes targeting the whole transcriptome and incubated the following day with immunofluorescent antibodies to define cell-type-enriched compartments (*PanCK*, trophoblast marker; *CD68*, macrophage marker; *CD31*, endothelial marker).

Slides were loaded onto the GeoMx instrument, and four ROIs were selected per placenta within the villous parenchyma of the mid-placenta. Intervillous regions were excluded from the ROI selection such that only villous tissue was profiled, thereby minimizing capture of maternal CD68⁺ immune cells within the intervillous space and enriching for fetal Hofbauer cells (**Figure S4a)**. ROI characteristics (predominant villous type, intervillous macrophage density, and adjacency to intervillous fibrin) were scored by investigators blinded to study group and compared using Fisher’s exact test. Within each ROI, AOIs were segmented according to morphology marker expression. Oligonucleotide-tagged RNA probes were then released by UV photocleavage and collected for downstream library preparation. Libraries were prepared using Seq Code primers and quality-checked with Agilent Bioanalyzer. Next-generation sequencing was then performed using the Illumina NovaSeqX at the University of California, San Francisco Center for Advanced Technology.

### Quality Control and Normalization

FASTQ files were converted to digital count conversion (DCC) files using the *GeoMxNGSPipeline* desktop application. DCC files were subsequently imported into R using the *GeomxTools* framework, and phenotypic annotations were merged from the study metadata.

Quality control (QC) thresholds were applied to AOIs to ensure adequate sequencing depth and tissue representation (**Table S2**). Probe-level QC was then performed using negative control probes to estimate background signal and to identify outlier probes by Grubbs’ test (α = 0.01). Limits of quantification (LOQ) were defined for each AOI as the geometric mean of negative probe counts multiplied by the geometric standard deviation squared. Genes detected above LOQ in fewer than 10% of segments were excluded. Counts were normalized by the 75th percentile (Q3) of expression within each AOI and log_2_-transformed for downstream analyses. Gene symbols and annotations were obtained from the HUGO Gene Nomenclature Committee (HGNC) database. Pseudogenes and poorly annotated loci were excluded prior to downstream analysis, only retaining genes encoding protein products and annotated non-coding RNAs (including lncRNA, miRNA, snoRNA, and snRNA).

### Validation and deconvolution of morphology-defined segments

Transcriptional identity of morphology-defined segments was assessed by PCA, canonical lineage-marker expression, pairwise differential expression, and unsupervised hierarchical clustering. Canonical markers of trophoblast (*KRT7, GATA3, CGA, CYP19A1*), fetal endothelial cells (*PECAM1, VWF, CDH5, KDR, CD34*), and Hofbauer cells (*CD68, CD163, FOLR2, CD14, MRC1*) were evaluated across segments. Pairwise differential expression between segments was assessed using linear mixed-effects models accounting for repeated sampling within placentas and slide-level effects, with adjustment for gravidity. Unsupervised hierarchical clustering was performed using the 10% of genes with the highest coefficient of variation across AOIs, using Pearson distance and average linkage.

Cell-type composition within each compartment (trophoblast, Hofbauer cell, and fetal endothelial cell) was inferred using *SpatialDecon* (R package) with a human placental single-cell RNA-seq reference (dbGaP accession phs001886.v5.p1).^9,58^ The reference dataset was restricted to fetal-origin cells from placental villous and basal plate (PVBP) samples, excluding decidual and other cell types not expected within villous AOIs. Reference cell-type expression profiles were generated by aggregating transcriptomic signals across annotated cell populations and restricted to genes shared with the GeoMx dataset. Cell-type abundance estimates were computed for each compartment using gene expression patterns shared with the reference dataset. Estimated cell-type proportions were compared between groups within each segment using linear mixed-effects models of logit-transformed proportions, adjusted for gravidity with slide and placenta as nested random effects.

### Differential Gene Expression

Differential gene expression was evaluated using linear mixed-effects models implemented in the *mixedModelDE* function within the *GeomxTools* package in R.

Analyses were performed separately for each of the three segments, representing regions enriched for trophoblasts (PanCK), fetal Hofbauer cells (CD68), and fetal endothelial cells (CD31). Models assessed differential expression between infection and control samples, controlling for gravidity, slide-level batch effects, and correlation among multiple AOIs obtained from the same placenta.

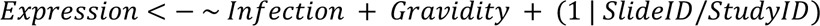

Statistical significance was defined as an absolute log_2_fold change ≥ 0.5 and a false discovery rate (FDR) < 0.20 using the Benjamini–Hochberg (BH) correction.

### Pathway Enrichment Analysis

Gene set enrichment analysis (GSEA) was performed using Gene Ontology (GO) gene sets for biological process (BP) using *clusterProfiler* (R).^59^ Gene sets containing fewer than 15 or greater than 500 genes were excluded. Genes were ranked according to a signed enrichment statistic *sign*(*β*) × −*log*₁₀*P*, with β representing the log_2_fold change and *P* representing the unadjusted p-value from the differential expression models. Enrichment *P* values were adjusted using the Benjamini–Hochberg method, and pathway enrichment was summarized using the normalized enrichment score (NES). Redundancy among GO Biological Process terms was reduced using the simplify function in *clusterProfiler*, with semantic similarity calculated using the Wang method (similarity cutoff = 0.4); among similar terms, the term with the lowest BH-adjusted *P* value was retained. Enriched pathways were visualized using the *enrichplot* and *ggplot2* packages in R.

Leading-edge genes were extracted from significantly enriched pathways identified by GSEA (FDR < 0.20). Candidate driver genes were defined as those present in the leading-edge subset and independently meeting significance criteria in the differential expression models (unadjusted *P* < 0.05 and |log_2_FC| ≥ 0.5).

As a complementary pathway-level analysis, Ingenuity Pathway Analysis (IPA; QIAGEN) Canonical Pathway Analysis was performed separately for each cell-enriched compartment. Genes with an unadjusted differential expression *P* value < 0.05 and |log_2_FC| ≥ 0.5 were included, together with their corresponding direction and magnitude of differential expression. Canonical pathway enrichment was assessed using IPA-generated unadjusted *P* values of overlap, with *P* < 0.05 considered significant. IPA activation z-scores were used to evaluate whether the observed direction of gene-expression changes was consistent with predicted pathway activation or inhibition based on curated molecular relationships; absolute z-scores ≥2 were considered evidence of predicted activation or inhibition.

### Histopathologic Assessment of Placental Lesions and Villous Extracellular Matrix Structure

To assess tissue-level correlates of the transcriptomic findings, placental specimens were selected for histopathologic and extracellular matrix analyses based on tissue availability. The Hematoxylin and Eosin (H&E) and Verhoeff’s Van Gieson (VVG) analyses were performed in partially overlapping cohorts. There were no significant differences in participant characteristics between cases and controls in either the H&E or VVG cohort (**Table S3**).

For histopathologic assessment, FFPE placental tissue from 8 cases with antepartum pyelonephritis and 15 controls was sectioned at 5 µm thickness and mounted onto glass slides. H&E staining was performed using standard protocols to evaluate placental villous architecture and histopathologic lesions. All H&E slides were reviewed by a subspecialty placental pathologist (C.G.) who was blinded to the study group. Placental lesions were characterized according to the Amsterdam Placental Workshop Group Consensus Statement and compared between cases and controls using Fisher’s exact test.^60^

To assess the extracellular matrix structure of the villous stroma, FFPE sections were VVG-stained according to the manufacturer’s instructions (Abcam*, ab150667*) and imaged with light microscopy at 20x (58 blocks from 30 participants: 8 pyelonephritis, 22 controls). Fetal arteries and veins in stem and intermediate villi were profiled separately (n = 1,790 vessels: 513 stem arteries, 398 intermediate arteries, 437 stem veins, and 442 intermediate veins). Terminal villi were excluded because of their limited extracellular matrix content. Each vessel was segmented into the lumen, the perivascular contractile sheath (PVCS) and the perivascular stroma (PVS). The PVCS was the inner fibromuscular ring of circumferentially arranged smooth muscle and elastic fibers surrounding the lumen. The PVS was the outer collagenous ring extending from the PVCS to the point where circumferentially oriented collagen fibers gave way to randomly oriented villous stroma.

After color normalization, pixels were classified (Trainable Weka Segmentation,^61^ FastRandomForest, Fiji/ImageJ) as collagen, elastin, nuclei or white space (representing background with no tissue). Vessel geometry (lumen diameter, PVCS and PVS thickness and area, and their ratios to lumen size) was also measured. Group differences were assessed with vessel-level linear mixed-effects models, with a random intercept for participant and adjustment for villous level. *P*-values are unadjusted. All 14 GeoMx participants also had VVG-stained slides. To relate transcriptomic and structural findings, participant-level *ELN* expression in each compartment was correlated with participant-level perivascular ECM composition (Spearman correlation). VVG image acquisition and analysis were performed with investigators blinded to study group.

## Data Availability

The data supporting the findings of this study are available from the corresponding author upon reasonable request.

---

**Table S1:** Definitions for Clinical Characteristics and Obstetric Outcomes.

| Variable | Definition |
| --- | --- |
| Pregestational BMI | Patient-reported BMI prior to pregnancy or earliest documented BMI during pregnancy |
| Pregestational diabetes mellitus | Diabetes diagnosed prior to pregnancy or hemoglobin A1C $\geq 6.5\%$ prior to 14 weeks' gestation |
| Gestational diabetes | Abnormal glucose tolerance testing after 14 weeks' gestation |
| Chronic hypertension | Elevated blood pressure diagnosed prior to pregnancy or before 20 weeks' gestation |
| Hypertensive disorder of pregnancy | New elevated blood pressure of 140/90mmHg or higher after 20 weeks' gestation |
| Fetal growth restriction | Sonographic estimated fetal weight or abdominal circumference $<10\%$ for gestational age by Hadlock criteria |
| Placental abruption | Clinical diagnosis based on vaginal bleeding, uterine findings, and/or gross placental findings with or without sonographic or pathologic confirmation. |
| Clinical chorioamnionitis | Intrapartum fever and maternal tachycardia, leukocytosis $>15,000$ per microliter, fetal tachycardia, purulent or foul-smelling amniotic fluid, or uterine tenderness. |
| Labor prior to delivery | Cervical dilation 1cm or greater, or ruptured membranes, prior to infant delivery |

**Table S2.** Quality control and filtering criteria for GeoMx data.

| Step | Metric | Threshold |
| --- | --- | --- |
| **Segment QC** | Total sequencing reads | $\geq 1,000$ |
| | Trimmed reads | $\geq 80\%$ |
| | Stitched reads | $\geq 80\%$ |
| | Aligned reads | $\geq 75\%$ |
| | Sequencing saturation | $\geq 50\%$ |
| | Negative control probe counts | $\geq 1$ |
| | No-template control counts | $\leq 9,000$ |
| | Nuclei per AOI | $\geq 20$ |
| | AOI area | $\geq 1,000 \mu\text{m}^2$ |
| **Probe QC** | Negative probe outliers | Grubbs' test, $\alpha = 0.01$ |
| | Probe ratio | $\geq 0.1$ |
| **Limit of quantification (LOQ)** | LOQ per AOI | Negative-probe geometric mean × (geometric SD)<sup>2</sup>; minimum = 2 |
| **Segment filtering** | Gene detection rate | $\geq 10\%$ of genes above LOQ |
| **Gene filtering** | Detection across AOIs | Above LOQ in $> 10\%$ of AOIs |
|  | Gene annotation | Current HGNC symbol with retained locus type |
| **Normalization** | Q3 normalization | 75th percentile |

**Table S3.** Characteristics of participants in H&E and Verhoeff-Van Gieson (VVG) studies.

|  | H&E |  |  | VVG |  |  |
| --- | --- | --- | --- | --- | --- | --- |
| Variable | Pyelonephritis (n=8) | Controls (n=15) | P-value | Pyelonephritis (n=8) | Controls (n=22) | P-value |
| **Maternal characteristics** |  |  |  |  |  |  |
| Maternal age – years, mean (SD) | 29.4 (7.8) | 32.9 (2.9) | 0.35 | 29.4 (7.8) | 32.9 (4.5) | 0.34 |
| Nulliparous | 6 (75.0) | 11 (73.3) | 1.00 | 6 (75.0) | 15 (68.2) | 1.00 |
| Obesity (BMI ≥30) | 1 (12.5) | 3 (20.0) | 1.00 | 1 (12.5) | 3 (13.6) | 1.00 |
| Chronic hypertension | 0 (0.0) | 0 (0.0) | N/A | 0 (0.0) | 0 (0.0) | N/A |
| Pregestational diabetes | 0 (0.0) | 0 (0.0) | N/A | 0 (0.0) | 0 (0.0) | N/A |
| **Obstetric characteristics** |  |  |  |  |  |  |
| Female fetus | 2 (25.0) | 6 (40.0) | 0.66 | 2 (25.0) | 10 (45.5) | 0.42 |
| Gestational age at delivery – weeks, mean (SD) | 38.7 (1.0) | 39.0 (1.1) | 0.35 | 38.7 (1.0) | 38.9 (1.1) | 0.67 |
| Preterm birth (<37 weeks) | 1 (12.5) | 1 (6.7) | 1.00 | 1 (12.5) | 1 (4.5) | 0.47 |
| Hypertensive disorder of pregnancy | 1 (12.5) | 1 (6.7) | 1.00 | 1 (12.5) | 3 (13.6) | 1.00 |
| Gestational diabetes | 1 (12.5) | 1 (6.7) | 1.00 | 1 (12.5) | 1 (4.5) | 0.47 |
| Fetal growth restriction | 1 (12.5) | 1 (6.7) | 1.00 | 1 (12.5) | 1 (4.5) | 0.47 |
| Placental abruption | 1 (12.5) | 1 (6.7) | 1.00 | 1 (12.5) | 1 (4.5) | 0.47 |
| Clinical chorioamnionitis | 2 (25.0) | 3 (20.0) | 1.00 | 2 (25.0) | 3 (13.6) | 0.59 |
| Labor prior to delivery | 7 (87.5) | 12 (80.0) | 1.00 | 7 (87.5) | 18 (81.8) | 1.00 |
| Vaginal delivery | 4 (50.0) | 10 (66.7) | 0.66 | 4 (50.0) | 15 (68.2) | 0.42 |

Data are presented as n (%) for categorical variables and mean (SD) for continuous variables. P-values were calculated using the Fisher's exact test for categorical variables and the Wilcoxon rank-sum test for continuous variables. BMI, body mass index (kg/m<sup>2</sup>).

**Figure S1.**
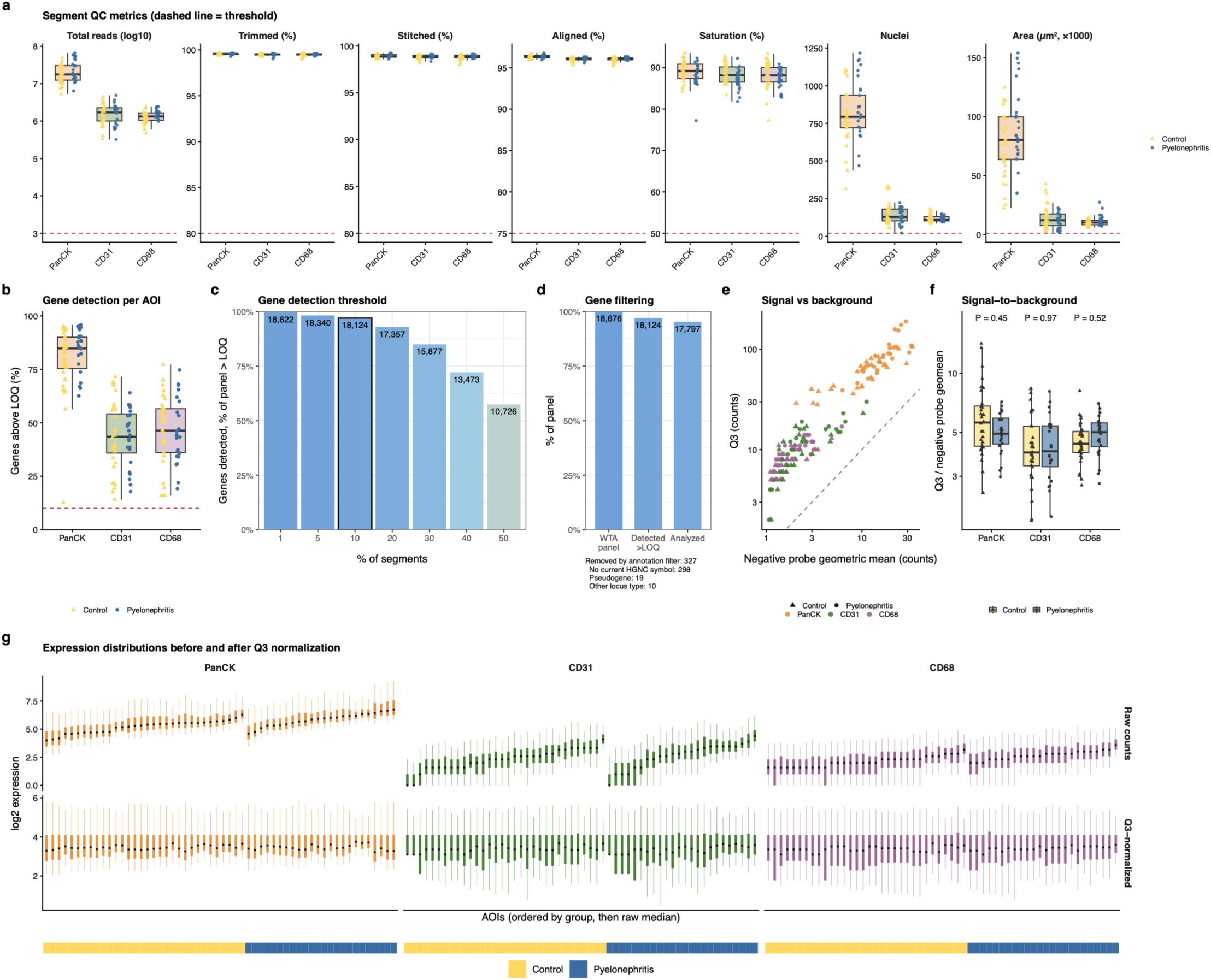
Quality control, gene filtering, and normalization of GeoMx data. **a,** Segment-level QC metrics for each AOI. Dashed red lines indicate predefined QC thresholds (**Table S2**); all AOIs met inclusion criteria. **b,** Percentage of WTA genes detected above the limit of quantification (LOQ) in each AOI; the dashed line indicates the 10% AOI inclusion threshold. **c,** Number of genes detected above the LOQ in more than the indicated percentage of AOIs. The outlined bar indicates the applied threshold (>10%). **d,** Genes retained after detection and annotation filtering. Excluded genes are summarized below the plot. **e,** Raw Q3 counts versus raw negative-probe geometric mean for each AOI, prior to normalization (log scale); the dashed line indicates where Q3 counts equal negative-probe counts. **f,** Signal-to-background ratio (raw Q3/negative-probe geometric mean) by segment and study group. P values are from linear mixed-effects models of log_2_-transformed ratios with placenta as a random effect. **g,** Distribution of log_2_ gene expression before (top) and after (bottom) Q3 normalization. Points indicate medians, thick bars interquartile ranges, and thin lines 5th–95th percentiles. AOIs are grouped by segment and study group; bars below indicate study group.

**Figure S2.**
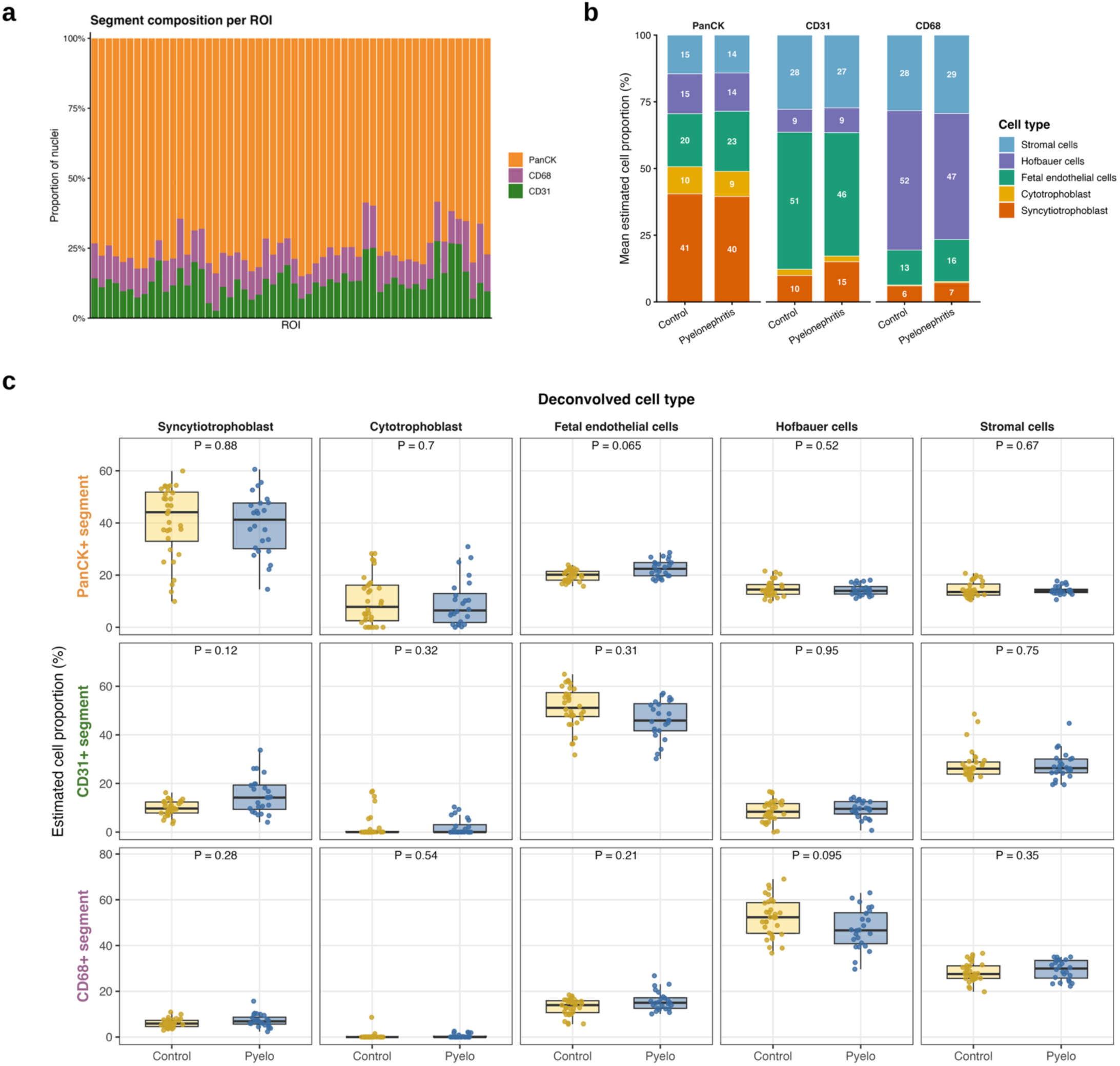
Estimated cell-type composition by study group. **a,** Relative contribution of PanCK-, CD68-, and CD31-segmented areas to the total nuclei count within each of the 56 ROIs. Each bar represents one ROI and is normalized to the total nuclei count across its three corresponding AOIs. PanCK-segmented areas accounted for the majority of nuclei across all ROIs. **b,** Mean *SpatialDecon*-estimated cell-type composition of PanCK-, CD31-, and CD68-segmented AOIs in control and pyelonephritis placentas. Stacked bars show the mean relative proportions of the deconvolved cell types within each morphology-defined segment and study group. **c,** *SpatialDecon*-estimated proportions of each deconvolved cell type within each morphology-defined segment, compared between control and pyelonephritis placentas. Points represent individual AOIs; boxes show the median and interquartile range. *P* values are unadjusted values from linear mixed-effects models of logit-transformed proportions, adjusted for gravidity, with slide and placenta as nested random effects. No significant differences in estimated cell-type composition were detected between control and pyelonephritis placentas, indicating that the observed transcriptional differences were not attributable to differences in cell-type composition.

**Figure S3.**
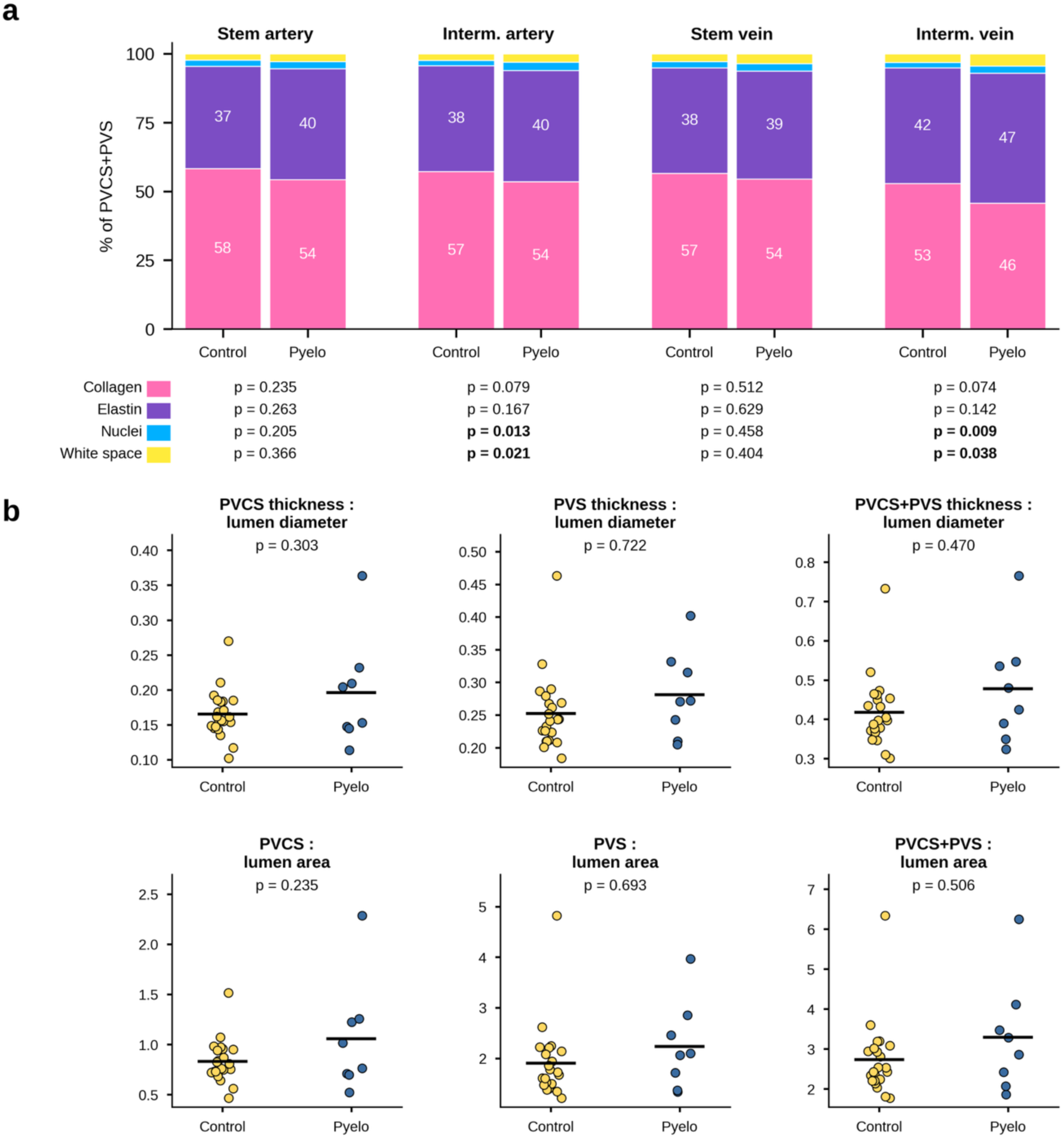
Perivascular ECM composition across villous vessel subtypes and vessel geometry. **a,** Mean composition of the combined perivascular zone (PVCS+PVS), stratified by study group and vessel subtype. Bars show the relative proportions of collagen, elastin, nuclei, and white space. *P* values are from linear mixed-effects models. **b,** Morphometric assessment of perivascular regions, including PVCS, PVS, and combined PVCS+PVS thickness and area normalized to lumen area. Each point represents one participant, with vessel measurements averaged within each placental FFPE block and then across blocks. Horizontal bars indicate group means. *P* values are from linear mixed-effects models, adjusted for villous level, with participant as a random intercept. No significant differences in perivascular geometry were observed between control and pyelonephritis placentas.

**Figure S4.**
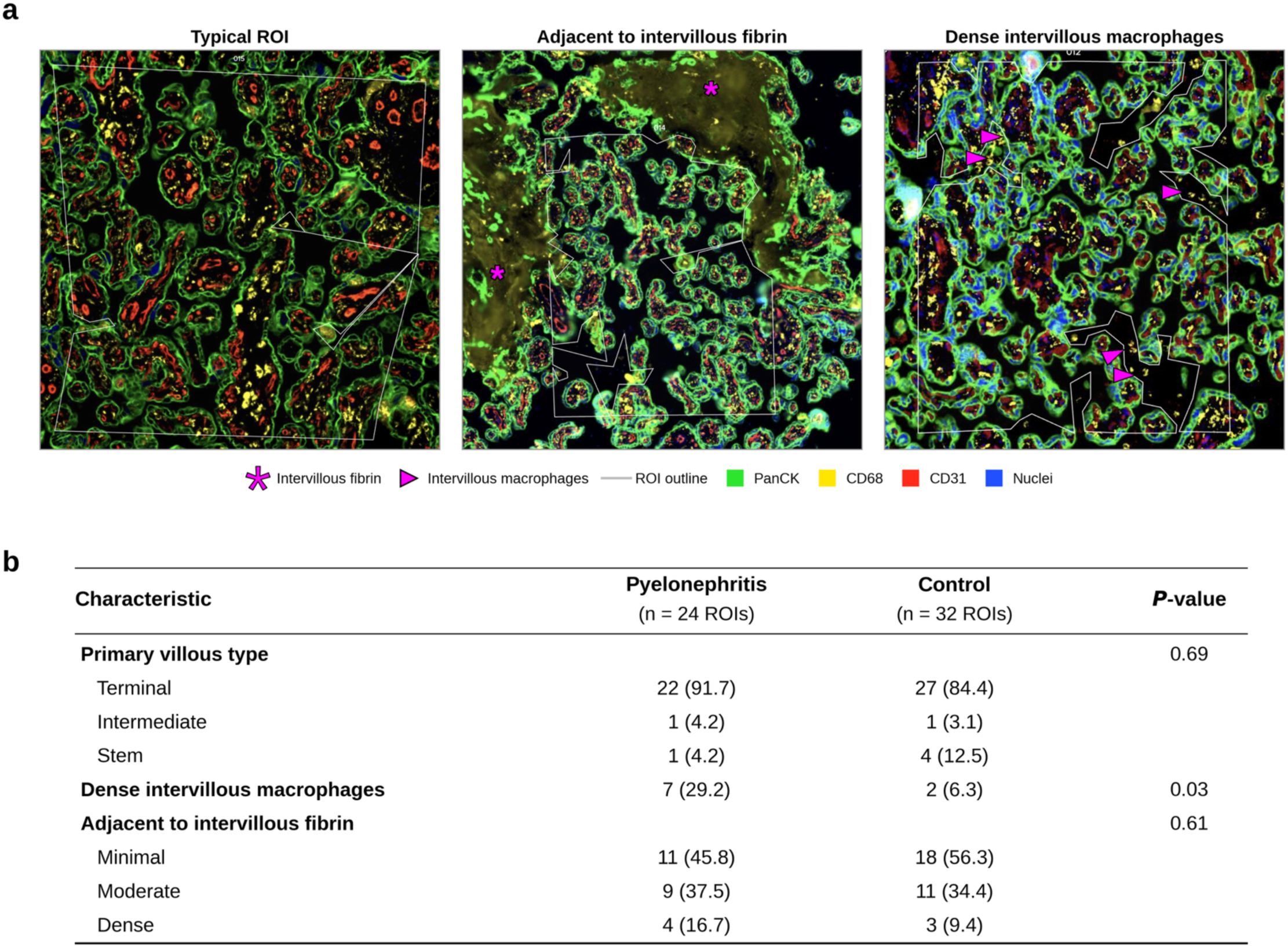
Histologic characteristics of selected GeoMx ROIs. **a,** Representative GeoMx DSP immunofluorescence images showing a typical ROI (left), an ROI adjacent to dense intervillous fibrin (middle) and an ROI with dense intervillous macrophages (right). White lines outline the ROIs. Magenta asterisks mark intervillous fibrin; magenta arrowheads mark clusters of CD68+ intervillous macrophages. **b,** ROI characteristics by study group, scored blinded to group on GeoMx immunofluorescence images. Primary villous type denotes the predominant villous morphology within each ROI; intervillous macrophage density and adjacency to intervillous fibrin were visually graded as categorical features. Values are n (%) of ROIs. *P* values are from Fisher’s exact test.

